# Ultrasonographic Splenic Indices Among Paediatric and Adults with Sickle Cell Disease in Nigeria

**DOI:** 10.1101/2023.02.24.23286418

**Authors:** Adama I Ladu, Caroline Jeffery, Abubakar Farate, Abubakar G Farouk, Fatima M Abulfathi, Adekunle Adekile, Imelda Bates

## Abstract

**Background:** Ultrasonography is an established and reliable method for assessing the spleen. Because of variation due to genetic and other environmental factors including malaria endemicity, interpretation of splenic sizes requires a knowledge of the normal reference range for a given population. The aim of this study was to determine spleen size in different age groups among healthy people in North-Eastern Nigeria and use this as a reference to determine spleen size amongst sickle cell disease (SCD) patients.

**Methods:** Using a cross-sectional study design, spleen size was measured in healthy people of different age groups, and steady-state SCD patients (children and adults) using abdominal ultrasonography. Using the age-group specific reference values obtained from the controls, spleens were classified into small, normal size, or enlarged among the SCD patients.

**Results:** Abdominal ultrasonography was performed for 313 participants, comprising 109 (34.8%) healthy controls and 204 (65.2%) steady-state SCD patients. The spleen was visualized in all the controls. However, 97(47.6%) of the SCD patients had no visible spleen. Small, normal, and enlarged spleens were observed in 16.7% (n=18/107), 63.6% (n=68/107) and 19.6% (n=21/107) SCD patients, respectively. Compared to the control group, splenic length was three-fold higher in the first two years of life in SCD patients, followed by a progressive age-related decline in size. Enlarged spleens were detected among 5(2.4%) SCD patients by manual palpation method compared to 21 (19.6%) using ultrasonography.

**Conclusion:** Model-based age-specific reference ranges and percentile curves for splenic dimensions based on ultrasonography among normal controls in North-Eastern Nigeria were established and may be of value in assessing spleen sizes among SCD patients living in malaria-endemic regions of Africa. Regular spleen scans to assess changes in size can help identify SCD patients at risk of splenomegaly complications including subclinical acute sequestration and hypersplenism, and those who are developing splenic atrophy.

## Introduction

Sickle cell disease (SCD) is an inherited condition of red blood cells widely prevalent across Sub-Saharan Africa, affecting up to 3% of births in some parts of the continent (1). The spleen is the largest organ in the reticulo-endothelial system with an active role in immune defence against infection and is one of the earliest organs to be affected in patients with SCD. In SCD the spleen initially enlarges, followed by progressive atrophy due to repeated episodes of vaso-occlusion and infarctions (2). Acute splenic sequestration crisis (ASSC), characterized by a sudden enlargement of the spleen can cause an abrupt drop in haemoglobin by at least 20% from baseline value, resulting in death in severe cases (3). Part of the routine clinical evaluation of patients with SCD involves examining for the spleen (4). However, clinical examination by palpation has a low sensitivity for the detection of splenic enlargement because the organ would have to enlarge two to three times its normal size before becoming palpable on abdominal examination (5); consequently, subclinical splenic sequestration crisis could go undetected during abdominal examination.

Ultrasonography provides a non-invasive method of assessing the spleen size; it is the test of choice in most climes because of its accuracy, low cost, flexibility, and safety profile (6). Other diagnostic modalities used in the radiological assessment of the spleen include CT and MRI scan (7-9), however, routine use of CT or MRI imaging is not feasible in most developing countries because of high cost and limited availability. Despite the widespread use of ultrasonography in the clinical evaluation of splenic enlargement, there is no consensus on how to define splenomegaly among SCD patients, as various cut-off values have been used in the literature (10-13). Some studies have considered a spleen length of ≥13 cm to indicate enlarged spleens (10, 11, 14, 15), while others have defined splenic enlargement as a spleen length of > 11cm (12) or > 12 cm (13). Some authors have used spleen length to define splenomegaly (16, 17), while others have used spleen volume (18, 19).

Furthermore, prior knowledge of the normal size of the spleen in a healthy population is required to interpret changes in spleen size because geographical variations in spleen size due to environmental factors like infections (e.g., malaria) and genetic variation can occur. Thus, establishing a reference range for populations living in the same area as the patients population is needed to be able to interpret ultrasonography assessments; however, few studies are available on normal spleen dimensions in children and adults in Africa (20-22), particularly those living in malaria-endemic regions (20-22).

Therefore, the aim of this study was to determine various splenic dimensions for different age groups in apparently healthy children and adults in North-Eastern Nigeria to generate a reference range for normal spleen size. This served as comparison for assessing changes in spleen size in patients with SCD in a tertiary hospital. We have also compared the spleen length and volume to assess spleen size, in order to identify which parameter is preferable and practical to reliably diagnose and follow up patients with splenomegaly in a resource-limited setting.

## Materials and methods

### Study design and participants

This hospital-based, cross-sectional study was conducted from October 2020 to November 2021 at the University of Maiduguri Teaching Hospital (UMTH), North-Eastern Nigeria. Healthy individuals consisting of medical students, children of hospital personnel, and post-op paediatric patients (without any acute or chronic illness likely to influence splenic size) on follow-up in the surgical clinic were enrolled into the study to establish spleen size reference ranges. Steady-state SCD patients (children and adults) on follow-up at the outpatient paediatric and haematology clinics during the study period were invited to take part in the study and their spleen sizes were compared to the reference ranges.

### Data collection

#### Clinical examination

Sociodemographic details of the study participants were collected using a structured questionnaire. Height (cm) and weight (kg) were measured, and body mass index (BMI) calculated as weight (kilograms) divided by body height (meters) squared. The spleen size was assessed by palpating the anterior axillary and mid clavicular line by a single examiner. Palpable splenomegaly was reported as the distance in cm that it extended under the left coastal margin in the mid clavicular line.

#### Sonographic evaluation

A board-certified radiologist with more than 15 years of experience in abdominal sonography performed all the examinations using Logic P5 Premium BT11 ultrasound scanner (GE Medical Systems, USA) equipped with a low frequency (3-5MHz) curvilinear transducer. The oblique intercostal approach was adopted for all patients and controls because this view provides an optimal window for good visualization of the spleen in most patients. The measurements were taken on sections through the splenic hilum as a fixed reference point. Splenic length (SL), defined as the maximum distance between the dome and the tip of the spleen, was measured in a longitudinal view. Splenic width (SW), defined as the maximum distance between the medial and lateral borders, was measured in a transverse view. Splenic depth (SD), defined as the maximum anteroposterior dimension, was also measured on a transverse view. The splenic volume was determined from these measurements using the standard prolate ellipsoid formula, SL x SW x SD × 0.523 (23). Two sequential measurements of each splenic parameter were obtained and the averages of the two measurements were used for statistical analysis. The spleen size of the SCD patients were compared with control values stratified by age. The spleen size was considered normal if the value fell within the 2.5^th^ and 97.5^th^ centile of the expected range for the age-specific group. Spleen sizes below and above the cut-off ranges were considered small-sized spleens or enlarged respectively (9, 20).

#### Statistical analysis

The data was analysed using Statistical Package for the Social Sciences (SPSS) (version 25; SPSS, Chicago, IL, USA). Categorical data were summarised using frequency and proportions while continuous data were summarised using descriptive statistics. Spearman’s correlation was used to determine the relationship between age, weight, height, body mass index, and ultrasonographic indices of spleen size. Model-based age-specific reference ranges were computed with age modelled as fractional polynomials using MedCalc® v.20.114 (MedCalc Statistical Software Ltd, Ostend, Belgium) (24). Log-transformation of variables was applied before model fitting as needed. The model-based 2.5^th^, 10^th^, 50^th^, 90^th,^ and 97.5^th^ percentiles of the (log-transformed) variable were then plotted against age. To assess intra-operator agreement for the ultrasonography measurements, the intra-class correlation coefficient (95% CI) and the Bland & Altman limits of agreement (25) were computed for each splenic parameter. The level of significance was set at two-tailed P-value <0.05.

#### Ethical considerations

The study was carried out according to the Declaration of Helsinki. Written informed consent was obtained from the adults and parents/guardians of the paediatric participants. The study protocol was approved by the University of Maiduguri Teaching Hospital (UMTH/REC/20/606) and Liverpool School of Tropical Medicine (LSTM) (REC reference number: 20-010) Ethics Review Boards.

## Results

### Study population

A total of 111 apparently normal controls and 214 patients with SCD were enrolled in the study. Ultrasonography data were available for 109 (98%) of the controls and 204 (95.3%) of the SCD patients; the remaining 12 (2 controls and 10 SCD patients) failed to turn up for the scan. Characteristics of the study participants are presented in Table 1. The spleen was clinically palpable in five of the SCD patients (n=5/204; 2.4%), ranging from 2 to 10 cm below the left coastal margin, but in none of the controls.

**Table 1:** Characteristics of study participants.

| Parameter | Controls | SCD patients |  | P value |
| --- | --- | --- | --- | --- |
| No of Subjects | 109 | 204 |  |  |
| Age (years, range) | 14.5 (1.5 – 34) | 13.0 (1.1 – 45) |  | 0.900 |
| No (%) of subjects per group. |  |  |  |  |
| < 5 years | 21 (19.3) | 42 (20.6) |  | 0.923 |
| 5 - 9 years | 22 (20.1) | 35 (17.1) |  |  |
| 10 – 14years | 21 (19.3) | 42 (20.6) |  |  |
| 15 years and above | 45 (41.3) | 85 (41.7) |  |  |
| Gender, n , (%) |  |  |  |  |
| Male | 68 (62.4) | 101 (49.5) |  | 0.030^^ |
| Female | 41 (37.6) | 103 (50.5) |  |  |
| Weight (kg), median, range | 36.0 (9 -100) | 29.0 (8 - 94) |  | 0.006* |
| Height (cm), median, range | 145 (74 -190) | 141.5 (73 – 188) |  | 0.152 |
| BMI (kg/m<sup>2</sup>), median, range | 17.9 (11 -30.8) | 15.5 (8 -30.4) |  | 0.000* |

P value ^^ Chi square, \*Mann Whitney U test. BMI body mass index

### Reliability test for spleen ultrasonography and relationship with anthropometric parameters

There was good intra-observer agreement and reliability for repeated measurements of all the splenic parameters (S. Table 1). Among the normal controls, the splenic length and volume showed a strong correlation with age, weight, height, and body mass index, whereas the correlations between splenic dimensions and anthropometric parameters were not as high among SCD patients (S. Table 2).

### Reference interval and percentile curves for spleen dimensions and age among the controls

The spleen was visualised on ultrasonography in all the control participants. Their spleen dimensions by age group and non-parametric reference limits are shown in Table 2. Specific reference limits for age 1 to 36 years are shown in Figs. 1a–d. Among the controls, 105 (96.3%; n=105/109) had spleen length within the 2.5^th^ and 97.5^th^ percentile range, and 4 (3.7%; n=4/109) above the 97.5^th^ centile (three were within the age group less than five years and one in the age group 15 years and above).

**Table 2:** Splenic dimensions according to age group and non-parametric reference ranges among the normal controls (n=109)

| Parameter | Number (n) | Mean (SD) | Median | 2.5<sup>th</sup> centile | 97.5<sup>th</sup> centile |
| --- | --- | --- | --- | --- | --- |
| **Spleen length (cm)** |  |  |  |  |  |
| < 5 years | 21 | 6.3 (0.7) | 6.1 | 5.2 | 7.0 |
| 5 – 9 years | 22 | 6.8 (0.9) | 6.9 | 5.3 | 8.3 |
| 10 – 14 years | 21 | 8.0 (1.2) | 7.7 | 6.0 | 11.1 |
| 15 years and above | 45 | 9.0 (1.4) | 8.8 | 7.2 | 12.5 |
| **Spleen depth (cm)** |  |  |  |  |  |
| < 5 years | 21 | 6.3 (0.7) | 6.2 | 5.1 | 8.1 |
| 5 – 9 years | 22 | 7.1 (0.9) | 7.1 | 5.4 | 8.5 |
| 10 – 14 years | 21 | 8.1(1.3) | 7.7 | 6.2 | 10.7 |
| 15 years and above | 45 | 9.2 (1.5) | 8.9 | 6.9 | 12.4 |
| **Spleen width (cm)** |  |  |  |  |  |
| < 5 years | 21 | 2.6(0.3) | 2.6 | 2.1 | 3.1 |
| 5 – 9 years | 22 | 3.0 (0.5) | 3.0 | 2.2 | 3.8 |
| 10 – 14 years | 21 | 3.3 (0.4) | 3.2 | 2.6 | 4.0 |
| 15 years and above | 45 | 4.0 (0.8) | 3.9 | 2.9 | 5.8 |
| **Spleen vol (cm<sup>3</sup>)** |  |  |  |  |  |
| < 5 years |  |  |  |  |  |
| 5 – 9 years | 21 | 54.0(14.9) | 53.5 | 32.0 | 86.4 |
| 10 – 14 years | 22 | 76.9(27.3) | 78.6 | 33.0 | 133 |
| 15 years and above | 21 | 117(48.7) | 102.8 | 56.2 | 226 |
|  | 45 | 180.8(87) | 152.7 | 80.0 | 414.7 |

**Fig1:**
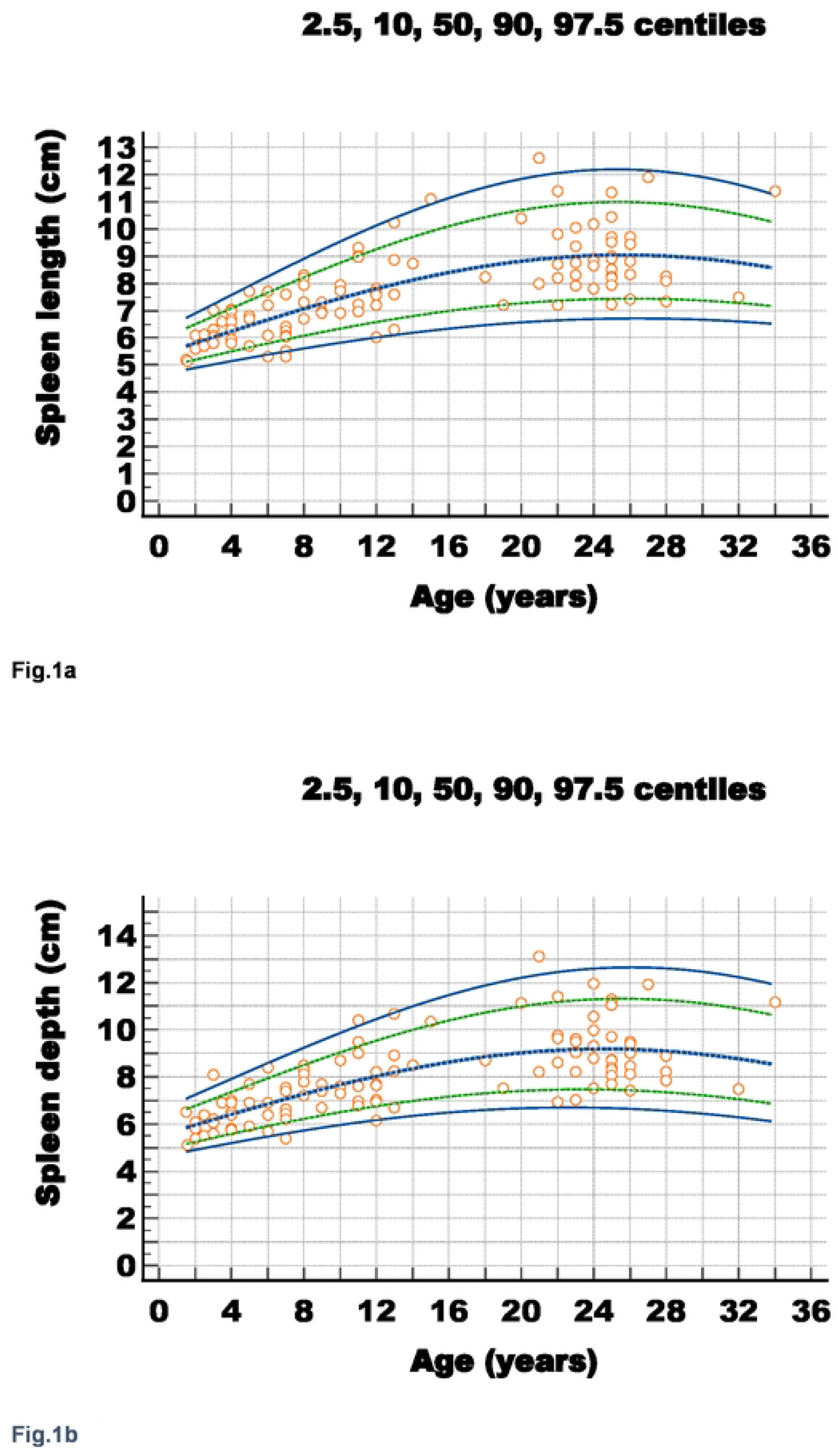
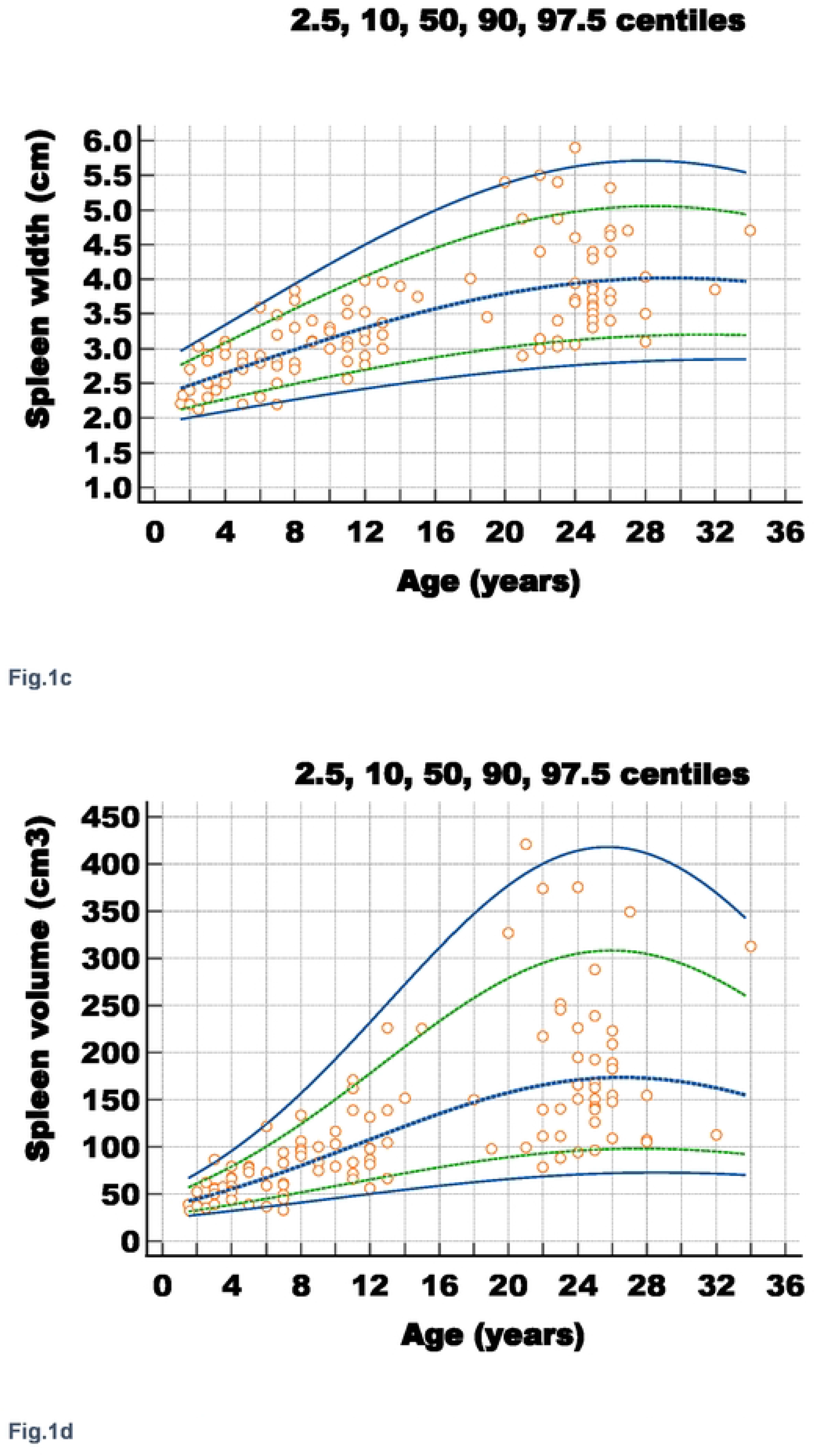
Percentile curves for various splenic dimensions. a: Percentile curve of the age-based reference limits for splenic length b: Percentile curve of the age-based reference limits for spleen depth c: Percentile curve of the age-based reference limits for spleen width d: Percentile curve of the age-based reference limits for spleen volume

### Spleen sizes among SCD patients

Of the 204 SCD patients, the spleen was visualized in 107 (52.3%) and not visualised in 97 (47.5%) by ultrasonography. Of the 107 patients with visible spleens, small, normal size and enlarged spleens occurred in 16.7% (n=18/107), 63.6% (n=68/107) and 19.6% (n=21/107) respectively, based on the reference range generated from our control participants. Further classification of spleen size across age groups is shown in Fig 2. and Table 3. Enlarged spleens were more frequently found in children less than five years old by both spleen length (31%) and volume (35.7%), and among patients 15 years above (25% and 20% by length and volume respectively). Small-sized spleens were more frequently encountered in the age group 10-14 years using either the spleen length (21.1%) or volume (31.6%) respectively (Table 3).

**Fig. 2:**
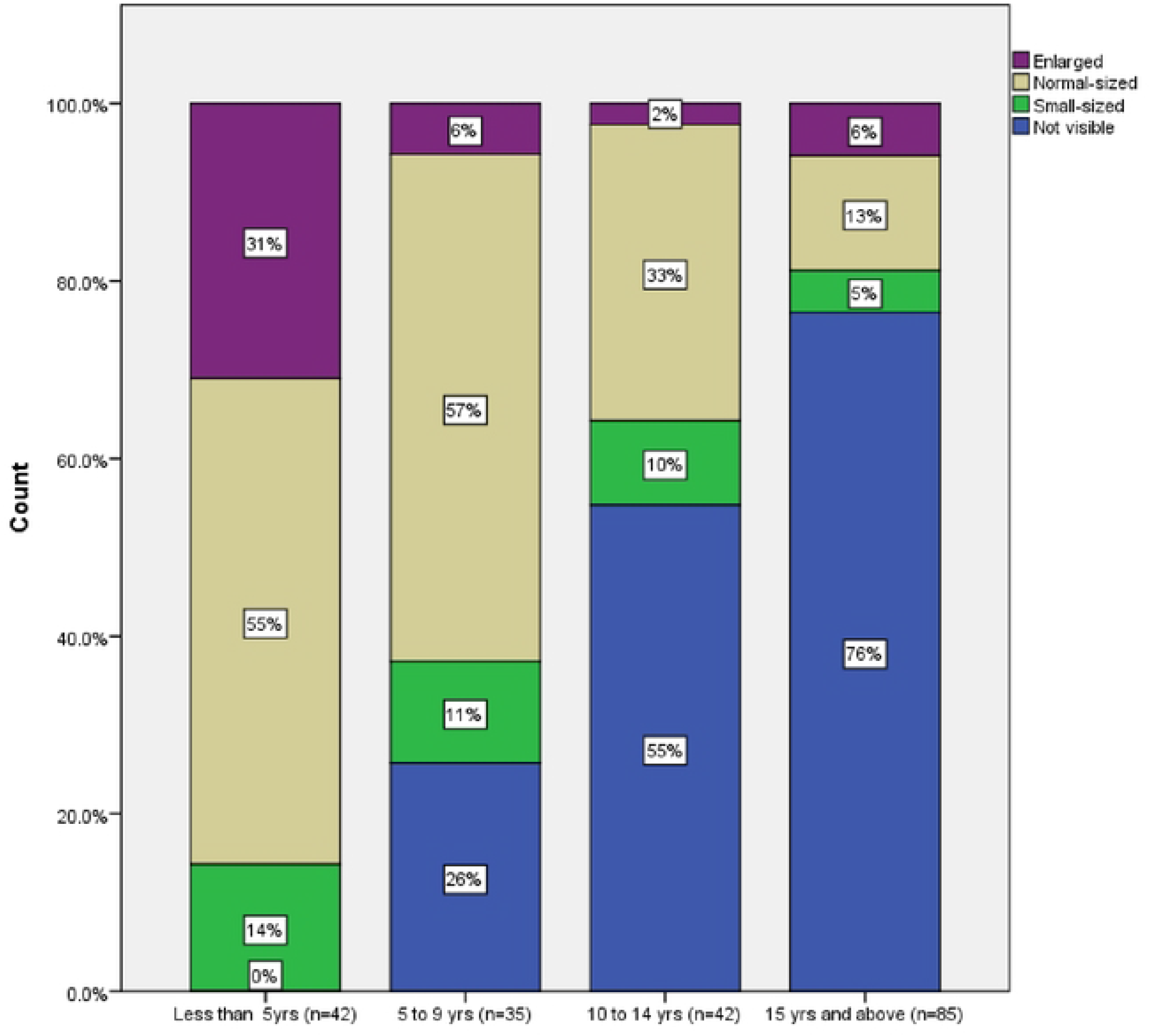
Distribution of spleen size according to age group. Distribution of spleen size among SCD patients (n=204) across different age groups assessed by ultrasonography. Spleen size was based on spleen length measurement.

**Table 3.** Frequency of different spleen sizes among SCD patients (n=107) by age group.

| Parameter | Reference interval |  | Category of spleen size, n, (%) |  |  | Total |
| --- | --- | --- | --- | --- | --- | --- |
|  | Lower limit<br>(2.5<sup>th</sup> centile) | Upper limit<br>(97.5<sup>th</sup> centile) | Small-sized | Normal | Enlarged |  |
| **Spleen length (cm)** |  |  |  |  |  |  |
| < 5 years | 5.0 | 7.0 | 6 (14.3) | 23 (54.8) | 13 (31.0) | 42 |
| 5 – 9 years | 5.5 | 8.5 | 4 (15.4) | 20 (76.9) | 2 (7.7) | 26 |
| 10 -14 years | 6.0 | 11.0 | 4 (21.1) | 14 (73.7) | 1 (5.3) | 19 |
| 15 years and above | 7.0 | 12.5 | 4 (20.0) | 11 (55.0) | 5 (25.0) | 20 |
| **Spleen volume (cm<sup>3</sup>)** |  |  |  |  |  |  |
| < 5 years | 32.0 | 87.0 | 4 (9.5) | 23 (54.8) | 15 (35.7) | 42 |
| 5 – 9 years | 33.0 | 133.0 | 2 (7.7) | 17 (65.4) | 7 (26.9) | 26 |
| 10 -14 years | 56.0 | 226.0 | 6 (31.6) | 12 (63.1) | 1 (5.3) | 19 |
| 15 years and above | 80.0 | 415.0 | 5 (25) | 11 (55.0) | 4 (20.0) | 20 |

Age-group based reference ranges were generated from the corresponding spleen length and volume of the controls. Spleen size below the 2.5<sup>th</sup> centile were considered small size; normal-sized spleen are dimensions between the 2.5<sup>th</sup> and 97.5<sup>th</sup> centiles; enlarged spleens are values above the 97.5<sup>th</sup> centiles.

### Comparison of spleen size and relationship with age between SCD patients and normal controls groups

Spleen length among the SCD patients with visible spleens was compared with those of normal controls. The mean spleen length (*P* = 0.000) and volume (*P* = 0.002) were significantly higher in the control group compared to SCD patients. This difference was particularly striking in those aged 10 – 14 years (Table 4). Of note however, the spleen length was almost three-fold higher among the SCD patients compared to the controls in the first two years of life, followed by a rapid decline in spleen size around the third year (Fig.3). Also, the fitted line between spleen size and age had a positive slope among different age groups of the control group (Fig. 4a), while the slope was negative across all age groups among the SCD patients (Fig.4b). When data for SCD patients with and without visible spleens were plotted, mixed positive and negative slopes were obtained across age groups (S.Fig.1).

**Table 4:** Comparison of spleen sizes between SCD patients and normal controls.

| Parameters | Sickle cell disease patients (n=107) |  |  | Normal controls (n=109) |  |  | P value |
| --- | --- | --- | --- | --- | --- | --- | --- |
|  | N | Mean(SD) | Median | N | Mean (SD) | Median |  |
| **Spleen length(cm)** |  |  |  |  |  |  |  |
| < 5 years | 42 | 6.4(1.2) | 6.5 | 21 | 6.2(0.6) | 6.1 | 0.423 |
| 5 – 9 years | 26 | 6.6(1.3) | 6.5 | 22 | 6.8(0.9) | 6.9 | 0.096 |
| 10 -14 years | 19 | 7.0(1.8) | 7.0 | 21 | 8.0(1.2) | 7.7 | 0.004* |
| >15 years | 20 | 9.9(3.6) | 8.8 | 45 | 9.0(1.3) | 8.8 | 0.921 |
| **Total** | 107 | 7.2 (2.3) | 6.7 | 109 | 7.8 (1.6) | 7.7 | 0.000* |
| **Spleen vol (cm<sup>3</sup>)** |  |  |  |  |  |  |  |
| < 5 years | 42 | 73.8(33.7) | 66.4 | 21 | 53.9 (14.6) | 53.5 | 0.020* |
| 5 – 9 years | 26 | 85.7(55.0) | 66.5 | 22 | 76.9 (28.0) | 78.6 | 0.979 |
| 10 -14 years | 19 | 90.0(60.0) | 70.4 | 21 | 117.0(48.3) | 102.0 | 0.013* |
| >15 years | 20 | 258.0(270) | 145.7 | 45 | 183.6(84.7) | 154.4 | 0.599 |
| **Total** | 107 | 114 (140) | 70.4 | 109 | 124 (80.8) | 99.5 | 0.002* |

\* P value: Mann Whitney U test for within age group analysis

**Fig. 3.** Relationship between spleen length and age among the less than five years participants. Scatter plot showing the relationship between spleen length (Y-axis) and age (X-axis) among the less than five years SCD patients (n=42) and normal controls (n=22). The spleen size was almost three-fold larger in the SCD population in the first two years of life compared to the controls, before decreasing abruptly around the third year; thereafter, the spleen size becomes variable as SCD with both small and large spleens could be observed. Regression lines are superimposed for both populations.

**Fig. 4.**
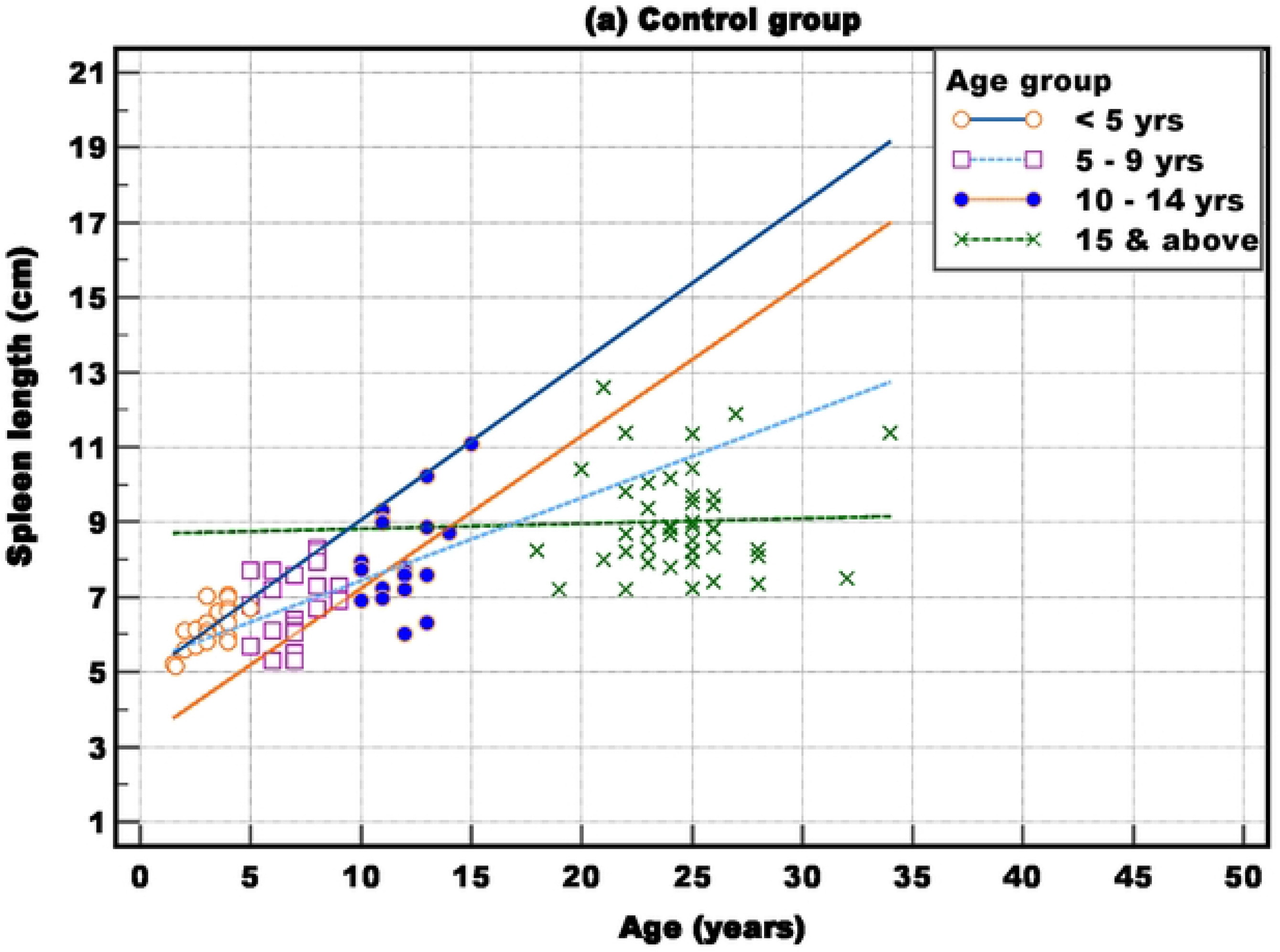
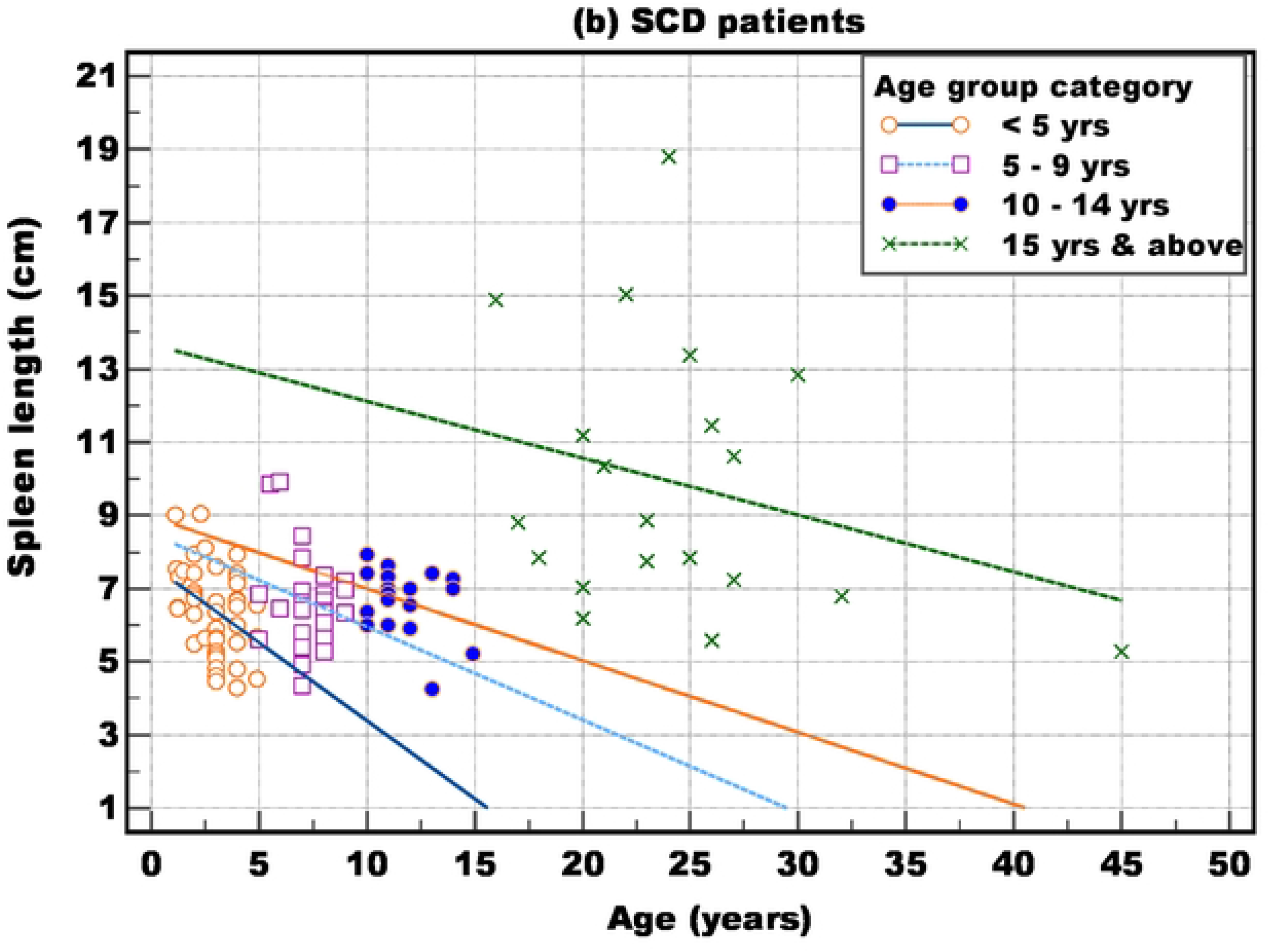
Relationship between spleen length and age. **a**: Scatter plot showing the relationship between spleen length (Y-axis) and age (X-axis) among the control group (n=109). The spleen length shows a progressive rise with increasing age up to the third decade before levelling off. Regression lines are superimposed for all age groups. **b**: Scatter plot showing the relationship between spleen length (Y-axis) and age (X-axis) among the SCD patients (n=107). The spleen length in the those less than five years (n=42) and those aged 5 to 9 years (n=26) were similar and both showed a downward trend. The spleen length becomes static in those aged 10 to 14 years (n=19) ; after 15 years the spleen length becomes variable with normal and enlarged spleens present (NB this represents the remaining 20 out of the 85 patients among this age group that still have their spleens visible). Regression lines superimposed for all age groups.

## Discussion

Ultrasonography is routinely used to evaluate the size of internal organs such as the spleen, because it is non-invasive, is not associated with radiation exposure, it provides real-time information and it is widely available in resource-limited settings (9); however, there are no standards for defining spleen size among SCD patients. Apart from the influence of age and geographical differences, variations in spleen sizes recorded in different studies can be attributed to differences in ultrasonography techniques and the parameters used to classify spleen size as normal or abnormal (26, 27). We documented splenic size and volume among apparently healthy participants from the same geographical area and used this as a reference for assessing spleen size among our SCD patients. We generated non-parametric (2.5^th^ – 97.5^th^ percentiles) reference ranges according to four age groups and smoothened curves to provide specific reference values for spleen sizes for individuals aged 1-36 years. The same radiologist obtained splenic measurements from all the study participants to ensure consistency and reduce inter-operator bias.

Differences in spleen sizes across race and geographic locations has been attributed to genetic and environmental factors including endemicity of infections associated with splenic enlargement. Our data on splenic length among the controls were similar to previous findings among paediatric (20-22) and adult population in Africa (6, 28, 29). In contrast, the upper limit of spleen length in our controls aged 1-4 years (n=22; median, 6.1 cm), 5-9 years (n=21; median 6.9 cm) and 10-14 years (n=22; median 7.7 cm) were smaller when compared among a normal population in the USA aged 2-4 years (n=24; median 7.4cm), 6-8 years (n=21; median 8.2 cm) and 10-12 years (n=17; median 9.9 cm) (27). Our data for the upper limit of spleen length were also smaller than those recorded among comparable age groups in Europe (9). In a study involving 631 American athletes, the spleen was larger in white Americans compared to the African-American athletes (30). These findings suggests that the spleen may be inherently smaller in the African population compared to the White population, contrary to the general notion of the spleen being bigger in areas where infections like malaria are endemic. Other studies from Nigeria have made similar observations of smaller spleen sizes among their population compared to published data among the White population (6, 28, 29). This underscores the importance of using population-specific reference values in classifying spleen sizes among individuals with SCD and other disease conditions affecting the spleen.

Studies on spleen size among SCD patients in Nigeria shows similarities within regions, and discordance across geographic regions. Whereas our findings of larger spleens in normal controls compared to the SCD patients concurs with findings from other parts of Northern Nigeria (11, 14, 16), it is in contrast with studies from South-West (19, 31) and South-East Nigeria (32, 33) where SCD patients were reported to have larger spleens than the control group. A previous study comparing spleen sizes among SCD patients across different regions in Nigeria, found that SCD patients from Southern Nigeria were more likely to have enlarged spleens (n=58/182; 31.9%) compared to those from the Northern part (n=11/128; 8.6%) (34). The reason for this difference was not clear; however, the authors surmised this could be related to the high rate of malaria transmission encountered in the rain forest belt of the South compared to the savanna grassland of the Northern part. Collaborative studies to investigate other modifying factors such as ethnicity and genetic factors between the two regions may be provide further insight.

Whereas most studies used spleen length to define splenomegaly in SCD (10-17, 36), others used spleen volume (18, 19, 37, 38) or both length and volume (31-33). Given the strong positive correlation between spleen length and volume in our normal controls (rho = 86.4; *P* = 0.0001), consistent with earlier reports (26, 39), both parameters were used to classify spleen sizes. A drawback of using spleen volume lies with the use of the prolate ellipsoid volume method in its determination formula. However, the spleen can become irregularly shaped in SCD patients due to the recurrent vaso-occlusion and infarctions, therefore, estimating volume using this formula may not be accurate (30). Despite similarities in the range of values obtained for the different categories of spleen sizes using spleen length and volume, we noted a tendency for higher frequencies of enlargement when using spleen volume. Furthermore, the ease of acquiring spleen length when compared to the cumbersome nature of assessing volume favours the use of spleen length to determine spleen size in routine clinical practice. More than half of our patients had their spleens visible on ultrasonography; we used the reference ranges generated from our controls to classify these spleens as normal-sized, small-sized or enlarged. About a third of the patients had spleen lengths within the normal range. Small-sized and enlarged spleens were observed in 8.8% and 10.0% of the SCD patients respectively. The majority of the controls had normal-sized spleens; only 3.4% had enlarged spleens and they were mostly among those less than five years of age. We also noted a difference in the pattern of age-related increase in spleen size between the controls and SCD patients. A good correlation between age and splenic length (rho = 0.763; *P* = 0.000) occurred among the controls; the splenic length showed a progressive increase in children less than five years to the adult mean size of 9.0 cm before levelling off, consistent with reports from the USA (27), Europe (9), and India (35). The SCD patients in our cohort had larger spleens early in life, but the rate of increase in length was slow thereafter. Compared to the progressive increase in length observed in the control group, the splenic length remained steady in SCD patients aged 5 – 9 years group, with a slight increase in the 10-14 years group, before increasing to the adult (>15 years) mean length of 9.9 cm. The slow increase in spleen size observed during childhood and adolescence among the SCD patients may be indicative of progressive splenic injury, thereby counteracting the normal age-related physiological increase in spleen size observed in the control group.

### Clinical implication

Although the spleen is usually relatively larger in infants and toddlers than in adults (9), we observed a more than 3-fold increase in size of the spleen among the SCD patients compared to the normal controls in the first two years of life. This is not unexpected as obstruction of the inter-endothelial slits in the basement membrane by the relatively rigid sickled red blood cells could result in passive splenic enlargement (40). Extra medullary haemopoiesis and infections may also account for enlarged spleens in early childhood among SCD patients (3, 41, 42). Our finding of enlarged spleen in the first two years of life, the age group most vulnerable to ASSC in SCD is clinically relevant. Although none of the patients in this study had a past history of ASSC or symptoms at the time of examination, in a retrospective cohort study of 190 cases, 75% of first episodes of ASSC occurred before 2 years of age (43). The diagnosis of ASSC is mainly clinical; however, ultrasonography may be helpful in identifying young SCD patients with splenomegaly, a group prone to acute sequestration thus requiring close monitoring. This is particularly important given that subtle splenic enlargement and ASSC could be missed on clinical examination (44). Although the majority (n=65/85) of our older SCD patients had no visible spleen on ultrasonography, the spleens among a quarter (n=5/20) of those with visualized spleens were found to be enlarged. It is not clear if the presence of compound heterozygosity for the Hb S gene may be associated with this finding (they consisted of 3 HbSS, 1 HbSC and 1 HbS thal). Patients with enlarged spleens may be prone to complication related to splenomegaly including chronic hypersplenism and may also need close monitoring.

The finding of enlarged spleens in 31% (n = 13/42) of our SCD patients less than five years of age using ultrasonography compared to 4.8% (n = 2/42) by clinical examination aptly demonstrates the low sensitivity of the latter technique and the fact that the spleen may be enlarged 2-3-fold before it becomes palpably enlarged. It is possible that red cell sequestration and hypersplenism occurs within a spleen that is not palpable and so splenic ultrasonography may be a useful adjunct to clinical management. The strong correlation among the splenic dimensions indicates that reliable measurements of the spleen can aid diagnosis and follow-up of this group of patients.

## Limitation

Our study has some limitations. Being a hospital-based and single centre study with small sample size used to generate the reference ranges may limit generalizability of the findings. Having a single board-certified radiologist to obtain all images in healthy participants and SCD group brought about consistency and improved accuracy of comparison between the groups, but this may not be possible in routine practice.

## Conclusion

This study has provided reference ranges and percentile curves for splenic dimensions for different ages based on ultrasonography among normal controls in North-Eastern Nigeria and may be of value in assessing spleen sizes among SCD patients living in malaria-endemic regions. The spleen length was three-fold higher in the first two years of life among the patients compared to controls, but this was followed by a progressive age-related decline in size suggesting progressive splenic injury. Regular spleen scans can help identify patients with enlarged spleens, who may require close monitoring for development of complications such as subclinical acute sequestration (especially in vulnerable age groups) and hypersplenism.

## Data Availability

All relevant data are within the manuscript and its Supporting Information files

NA

## Acknowledgement

The authors are grateful to staff of the Radiology and Haematology Departments of the UMTH for their assistance during the data collection, and ultrasonography. We also thank all the patients and their guardians.

## Authors’ contributions

AIL and IB conceived and designed the project. AIL, AGF and FMA coordinated the patient’s recruitment. AIL and AF performed the ultrasonography. AIL performed the statistical analysis. IB, CJ and AA supervised the formal analysis. All the authors contributed significant intellectual content during the manuscript drafting or revision and accept responsibility for the integrity of the data and the accuracy of the analysis.

